# Characteristics and outcomes of 627 044 COVID-19 patients with and without obesity in the United States, Spain, and the United Kingdom

**DOI:** 10.1101/2020.09.02.20185173

**Authors:** Martina Recalde, Elena Roel, Andrea Pistillo, Anthony G. Sena, Albert Prats-Uribe, Waheed-Ul-Rahman Ahmed, Heba Alghoul, Thamir M Alshammari, Osaid Alser, Carlos Areia, Edward Burn, Paula Casajust, Dalia Dawoud, Scott L. DuVall, Thomas Falconer, Sergio Fernández-Bertolín, Asieh Golozar, Mengchun Gong, Lana Yin Hui Lai, Jennifer C.E. Lane, Kristine E. Lynch, Michael E. Matheny, Paras P Mehta, Daniel R. Morales, Karthik Natarjan, Fredrik Nyberg, Jose D. Posada, Christian G. Reich, Lisa M. Schilling, Karishma Shah, Nigam H. Shah, Vignesh Subbian, Lin Zhang, Hong Zhu, Patrick Ryan, Daniel Prieto-Alhambra, Kristin Kostka, Talita Duarte-Salles

**Author notes:** Joint first authors (Martina Recalde, Elena Roel). Joint last authors (Kristin Kostka, Talita Duarte-Salles). **Corresponding author:** Talita Duarte-Salles, Fundació Institut Universitari per a la recerca a l’Atenció Primària de Salut Jordi Gol i Gurina (IDIAPJGol), Gran Via Corts Catalanes, 587 àtic, 08007 Barcelona - Spain.

## Abstract

**Background:** COVID-19 may differentially impact people with obesity. We aimed to describe and compare the demographics, comorbidities, and outcomes of obese patients with COVID-19 to those of non-obese patients with COVID-19, or obese patients with seasonal influenza.

**Methods:** We conducted a cohort study based on outpatient/inpatient care, and claims data from January to June 2020 from the US, Spain, and the UK. We used six databases standardized to the OMOP common data model. We defined two cohorts of patients *diagnosed* and/or *hospitalized* with COVID-19. We created corresponding cohorts for patients with influenza in 2017-2018. We followed patients from index date to 30 days or death. We report the frequency of socio-demographics, prior comorbidities, and 30-days outcomes (hospitalization, events, and death) by obesity status.

**Findings:** We included 627 044 COVID-19 (US: 502 650, Spain: 122 058, UK: 2336) and 4 549 568 influenza (US: 4 431 801, Spain: 115 224, UK: 2543) patients. The prevalence of obesity was higher among *hospitalized* COVID-19 (range: 38% to 54%) than *diagnosed* COVID-19 (30% to 47%), or *diagnosed* (15% to 47%) or *hospitalized* (27% to 48%) influenza patients. Obese *hospitalized* COVID-19 patients were more often female and younger than non-obese COVID-19 patients or obese influenza patients. Obese COVID-19 patients were more likely to have prior comorbidities, present with cardiovascular and respiratory events during hospitalization, require intensive services, or die compared to non-obese COVID-19 patients. Obese COVID-19 patients were more likely to require intensive services or die compared to obese influenza patients, despite presenting with fewer comorbidities.

**Interpretation:** We show that obesity is more common amongst COVID-19 than influenza patients, and that obese patients present with more severe forms of COVID-19 with higher hospitalization, intensive services, and fatality than non-obese patients. These data are instrumental for guiding preventive strategies of COVID-19 infection and complications.

**Funding:** The European Health Data & Evidence Network has received funding from the Innovative Medicines Initiative 2 Joint Undertaking (JU) under grant agreement No 806968. The JU receives support from the European Union’s Horizon 2020 research and innovation programme and EFPIA. This research received partial support from the National Institute for Health Research (NIHR) Oxford Biomedical Research Centre (BRC), US National Institutes of Health, US Department of Veterans Affairs, Janssen Research & Development, and IQVIA. The University of Oxford received funding related to this work from the Bill & Melinda Gates Foundation (Investment ID INV-016201 and INV-019257). APU has received funding from the Medical Research Council (MRC) [MR/K501256/1, MR/N013468/1] and Fundación Alfonso Martín Escudero (FAME) (APU). VINCI [VA HSR RES 13-457] (SLD, MEM, KEL). JCEL has received funding from the Medical Research Council (MR/K501256/1) and Versus Arthritis (21605). No funders had a direct role in this study. The views and opinions expressed are those of the authors and do not necessarily reflect those of the Clinician Scientist Award programme, NIHR, Department of Veterans Affairs or the United States Government, NHS, or the Department of Health, England.

**Research in context:** *Evidence before this study:* Previous evidence suggests that obese individuals are a high risk population for COVID-19 infection and complications. We searched PubMed for articles published from December 2019 until June 2020, using terms referring to SARS-CoV-2 or COVID-19 combined with terms for obesity. Few studies reported obesity and most of them were limited by small sample sizes and restricted to hospitalized patients. Further, they used different definitions for obesity (i.e. some reported together overweight and obesity, others only reported obesity with BMI>40kg/m^2^). To date, no study has provided detailed information on the characteristics of obese COVID-19 patients, such as the prevalence of comorbidities or COVID-19 related outcomes. In addition, despite the fact that COVID-19 has been often compared to seasonal influenza, there are no studies assessing whether obese patients with COVID-19 differ from obese patients with seasonal influenza.

*Added value of this study:* We report the largest cohort of obese patients with COVID-19 and provide information on more than 29 000 aggregate characteristics publicly available. Our findings were consistent across the participating databases and countries. We found that the prevalence of obesity is higher among COVID-19 compared to seasonal influenza patients. Obese patients with COVID-19 are more commonly female and have worse outcomes than non-obese patients. Further, they have worse outcomes than obese patients with influenza, despite presenting with fewer comorbidities.

*Implications of all the available evidence:* Our results show that individuals with obesity present more comorbidities and worse outcomes for COVID-19 than non-obese patients. These findings may be useful in guiding clinical practice and future preventative strategies for obese individuals, as well as provide useful data to support subsequent association studies focussed on obesity and COVID-19.

## Introduction

Obesity is associated with increased mortality and is a well-known risk factor of chronic conditions, such as diabetes, hypertension, cardiovascular disease and, cancer.^1^ Due to its proinflammatory state that impairs the immune response, obesity has also been related to increased risk of viral infections, including seasonal influenza.^2^

The novel coronavirus disease 2019 (COVID-19), caused by the severe acute respiratory syndrome coronavirus 2 (SARS-CoV-2), has been compared to seasonal influenza in terms of symptoms and complications.^3^ Both viruses cause respiratory tract infection with clinical manifestations ranging from asymptomatic/mild symptoms to severe illness requiring intensive services. Partly due to its similarities with influenza, people with obesity were soon labelled as “at-risk” individuals.^4^ Recent studies have found that obesity is common among severe and fatal COVID-19 cases.^5-8^ In hospitalized cohorts from the United States (US), obesity prevalence in COVID-19 cases ranges from 40 to 50%,^9-11^ while lower prevalence has been reported in non-hospitalized cases. A primary care study from Spain found that 20% of COVID-19 cases were obese,^12^ while in a population-based study from Denmark only 9% were obese.^13^

Since obesity is a worldwide public health priority, granular information on obese patients with COVID-19 is needed to guide preventive strategies.^14^ In this study, we aimed to describe and compare the demographics, comorbidities, and outcomes of obese patients with COVID-19 to those of 1) non-obese patients with COVID-19, and 2) obese patients with seasonal influenza, among inpatient or outpatient settings.

## Methods

### Study design, setting and data sources

We conducted a multinational cohort study using routinely-collected healthcare data from January to June 2020 from the US, Spain, and the United Kingdom (UK). All data were standardized to the Observational Medical Outcomes Partnership (OMOP) Common Data Model (CDM).^15^ The open-science Observational Health Data Sciences and Informatics (OHDSI) network maintains the OMOP-CDM, along with a wide range of tools developed by its members to facilitate analyses of mapped data.^16^

We included primary, outpatient and inpatient care data from electronic health records (EHRs) and health insurance claims data. Primary care data sources included the Clinical Practice Research Datalink (CPRD), with patients from over 600 general practices in the UK;^17^ and the Information System for Research in Primary Care (SIDIAP), covering approximately 80% of the population in Catalonia, Spain.^18^ Data from the US included: the Stanford Medicine Research Data Repository (STARR-OMOP), with data from Stanford Health Care,^19^ Columbia University Irving Medical Center (CUIMC), covering New York-Presbyterian Hospital and its affiliated physician practices; IQVIA Open Claims, which are pre-adjudicated claims collected from office based physicians and specialists covering over 300 million lives (~80% of the US population); and the United States Department of Veterans Affairs (VA-OMOP), covering the national Department of Veterans Affairs health care system which serves more than 9 million enrolled Veterans (of whom 93% are male). A more detailed description of the included data sources is available in Appendix 1.

### Study participants

For our first objective (obese vs non-obese patients with COVID-19), we included two cohorts of patients: 1) all patients *diagnosed* with COVID-19 (clinical diagnosis and/or positive test for SARS-CoV-2), and, 2) all patients *hospitalized* with a COVID-19 diagnosis. The codes used to identify a COVID-19 diagnosis are described in Appendix 2. We included individuals with at least one year of observation time prior to index date to capture observed baseline characteristics. In the *diagnosed* cohort, index date was defined as the date of COVID-19 clinical diagnosis or the earliest test day registered within seven days of a first positive test, whichever occurred first. In the *hospitalized* cohort, index date was the day of hospitalization. Patients were followed from the index date to the earliest of death, end of the observation period,^20^ or 30 days.

Patients *hospitalized* with COVID-19 were identified as those having a hospitalization episode along with a clinical diagnosis or positive SARS-CoV-2 test within a time window from 21 days prior to admission up to the end of their hospitalization. We chose this time window to include patients with a diagnosis prior to hospitalization and to allow for a record delay in test results or diagnoses.

Both the *diagnosed* and *hospitalized* COVID-19 cohorts were stratified by obesity (obese vs non-obese). Obesity was defined as having an ever-recorded obesity diagnosis (Appendix 3) and/or a body mass index (BMI) measurement between 30 and 60 kg/m^2^ and/or a body weight measurement between 120 and 200 kilograms prior or at index date. Non-obese patients were those who did not fulfil the obesity definition.

For our second objective (obese COVID-19 vs obese influenza patients), we reproduced these cohorts for patients with a clinical diagnosis and/or a positive test for influenza in the season 2017-2018 (Appendix 2).

### Baseline characteristics and outcomes of interest

We obtained information on the participants’ sex (female, male) and age at index date. Clinical epidemiologists generated a list of codes for the identification of prior medical conditions and outcomes of interest using a web-based integrated platform (ATLAS tool).^20^

We identified comorbidities from up to one year prior to index date. We selected conditions based on their prevalence in the cohorts of the participating sites, as well as on their clinical relevance to the obesity and the COVID-19 research field.^4,21^ Comorbidities were identified based on the Systematized Nomenclature of Medicine (SNOMED) hierarchy, with all descendant codes included.^20^ We created specific definitions for comorbidities of particular interest; the detailed definitions of these variables can be consulted in Appendix 3.

Our main 30-day outcomes of interest were hospitalization and death for the *diagnosed* cohorts, and requirement of intensive services (identified by a recorded mechanical ventilation and/or a tracheostomy and/or extracorporeal membrane oxygenation procedure) and death for the *hospitalized* cohorts. For the *hospitalized* cohort of COVID-19 patients, we also report respiratory, cardiovascular, thromboembolic, and other events occurring in the 30 days after the index date.

### Statistical analysis

We describe the number of patients included and the prevalence of obesity in each database. We report the percentage of COVID-19 diagnoses that was identified by a positive SARS-CoV-2 test, as well as the socio-demographics, comorbidities, and outcomes as proportions (calculated by the number of persons within a given category, divided by the total number of persons) for each database, by obesity status.

A common analytical code was developed for the OHDSI Methods library which was run locally in each database (available at https://github.com/ohdsi-studies/Covid19CharacterizationCharybdis), and only aggregate results from each database were publicly-shared.

We used R version 3.6 for data visualization. All the data partners obtained Institutional Review Board (IRB) approval or exemption to conduct this study.

## Results

### Prevalence of obesity

The COVID-19 dataset included 627 044 *diagnosed* and 160 013 *hospitalized patients*. The *diagnosed* cohort consisted of 502 650 from the US (STARR-OMOP: 3328; CUIMC: 8519; IQVIA-OpenClaims: 466 191; VA-OMOP: 24 612), 122 058 from Spain (SIDIAP), and 2336 patients from the UK (CPRD). The *hospitalized* cohort included 141 816 patients from the US (STARR-OMOP: 615; CUIMC: 2600; IQVIA-OpenClaims: 133 091 and VA-OMOP: 5510) and 18 197 from Spain (SIDIAP). Among *diagnosed* and *hospitalized* patients, 207 859 (33%) and 63 866 (40%) were obese, respectively. In all databases, the prevalence of obesity was lower among *diagnosed* patients than among those *hospitalized*, with differences ranging from 5% (IQVIA-OpenClaims) to 16% (SIDIAP) (Table 1).

**Table 1:**
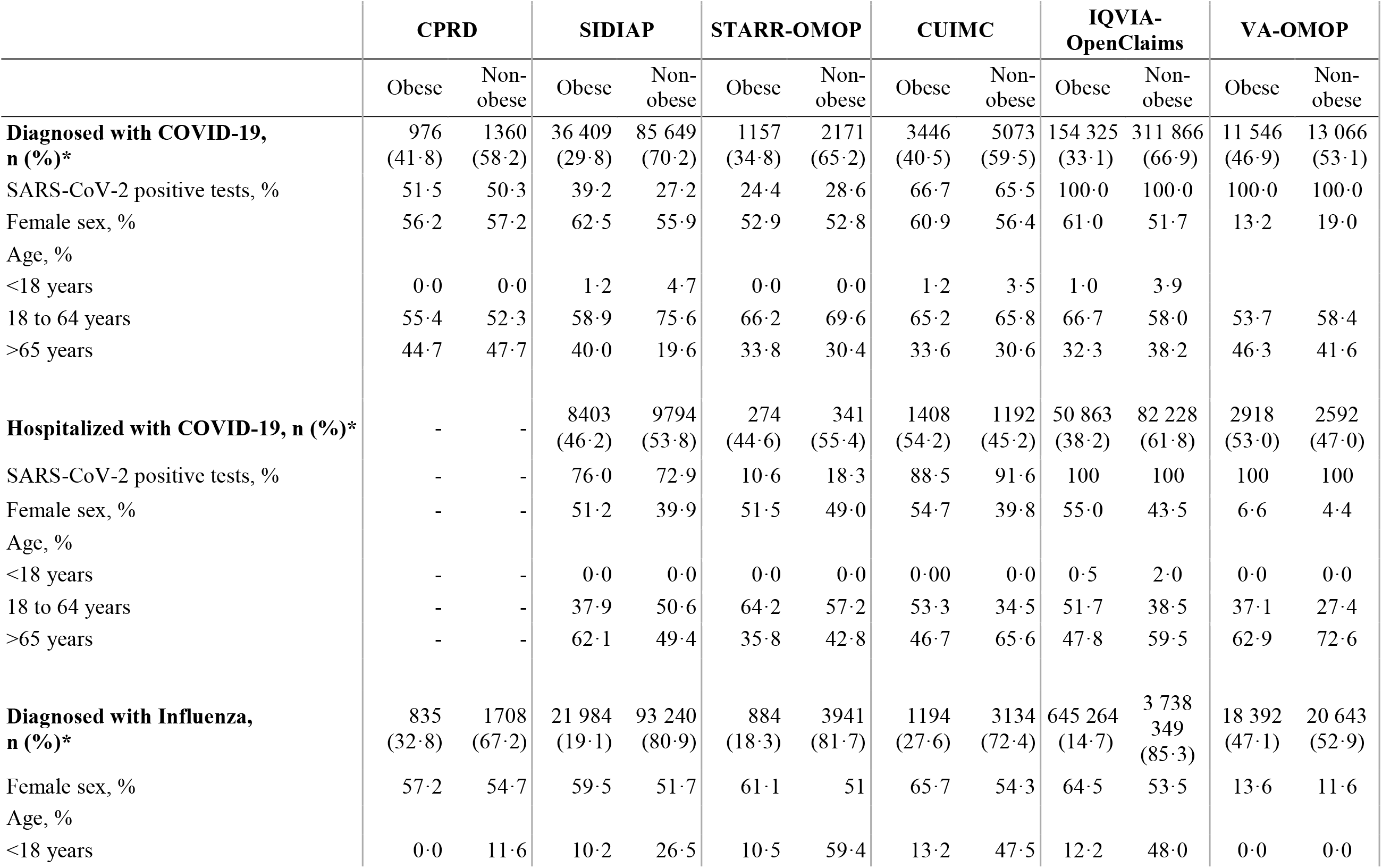
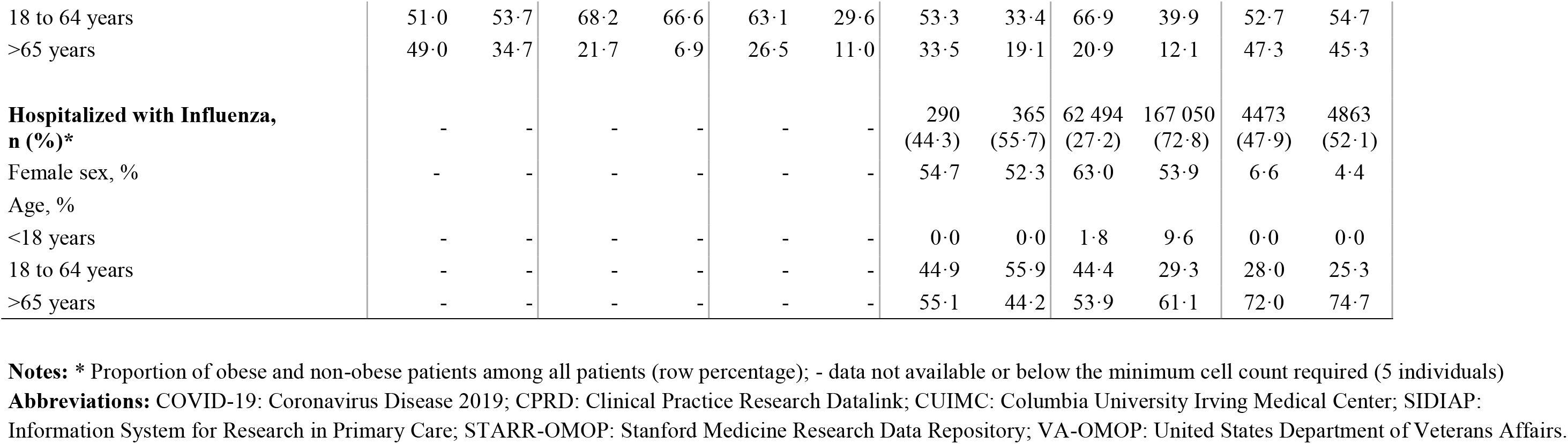
Sociodemographic characteristics of obese and non-obese patients diagnosed and hospitalized with COVID-19 and influenza in the season 2017-2018.

|  | CPRD |  | SIDIAP |  | STARR-OMOP |  | CUIMC |  | IQVIA-OpenClaims |  | VA-OMOP |  |
| --- | --- | --- | --- | --- | --- | --- | --- | --- | --- | --- | --- | --- |
|  | Obese | Non-obese | Obese | Non-obese | Obese | Non-obese | Obese | Non-obese | Obese | Non-obese | Obese | Non-obese |
| **Diagnosed with COVID-19, n (%)\*** | 976<br>(41.8) | 1360<br>(58.2) | 36 409<br>(29.8) | 85 649<br>(70.2) | 1157<br>(34.8) | 2171<br>(65.2) | 3446<br>(40.5) | 5073<br>(59.5) | 154 325<br>(33.1) | 311 866<br>(66.9) | 11 546<br>(46.9) | 13 066<br>(53.1) |
| SARS-CoV-2 positive tests, % | 51.5 | 50.3 | 39.2 | 27.2 | 24.4 | 28.6 | 66.7 | 65.5 | 100.0 | 100.0 | 100.0 | 100.0 |
| Female sex, % | 56.2 | 57.2 | 62.5 | 55.9 | 52.9 | 52.8 | 60.9 | 56.4 | 61.0 | 51.7 | 13.2 | 19.0 |
| Age, % |  |  |  |  |  |  |  |  |  |  |  |  |
| <18 years | 0.0 | 0.0 | 1.2 | 4.7 | 0.0 | 0.0 | 1.2 | 3.5 | 1.0 | 3.9 |  |  |
| 18 to 64 years | 55.4 | 52.3 | 58.9 | 75.6 | 66.2 | 69.6 | 65.2 | 65.8 | 66.7 | 58.0 | 53.7 | 58.4 |
| >65 years | 44.7 | 47.7 | 40.0 | 19.6 | 33.8 | 30.4 | 33.6 | 30.6 | 32.3 | 38.2 | 46.3 | 41.6 |
| **Hospitalized with COVID-19, n (%)\*** | - | - | 8403<br>(46.2) | 9794<br>(53.8) | 274<br>(44.6) | 341<br>(55.4) | 1408<br>(54.2) | 1192<br>(45.2) | 50 863<br>(38.2) | 82 228<br>(61.8) | 2918<br>(53.0) | 2592<br>(47.0) |
| SARS-CoV-2 positive tests, % | - | - | 76.0 | 72.9 | 10.6 | 18.3 | 88.5 | 91.6 | 100 | 100 | 100 | 100 |
| Female sex, % | - | - | 51.2 | 39.9 | 51.5 | 49.0 | 54.7 | 39.8 | 55.0 | 43.5 | 6.6 | 4.4 |
| Age, % |  |  |  |  |  |  |  |  |  |  |  |  |
| <18 years | - | - | 0.0 | 0.0 | 0.0 | 0.0 | 0.00 | 0.0 | 0.5 | 2.0 | 0.0 | 0.0 |
| 18 to 64 years | - | - | 37.9 | 50.6 | 64.2 | 57.2 | 53.3 | 34.5 | 51.7 | 38.5 | 37.1 | 27.4 |
| >65 years | - | - | 62.1 | 49.4 | 35.8 | 42.8 | 46.7 | 65.6 | 47.8 | 59.5 | 62.9 | 72.6 |
| **Diagnosed with Influenza, n (%)\*** | 835<br>(32.8) | 1708<br>(67.2) | 21 984<br>(19.1) | 93 240<br>(80.9) | 884<br>(18.3) | 3941<br>(81.7) | 1194<br>(27.6) | 3134<br>(72.4) | 645 264<br>(14.7) | 3 738 349<br>(85.3) | 18 392<br>(47.1) | 20 643<br>(52.9) |
| Female sex, % | 57.2 | 54.7 | 59.5 | 51.7 | 61.1 | 51 | 65.7 | 54.3 | 64.5 | 53.5 | 13.6 | 11.6 |
| Age, % |  |  |  |  |  |  |  |  |  |  |  |  |
| <18 years | 0.0 | 11.6 | 10.2 | 26.5 | 10.5 | 59.4 | 13.2 | 47.5 | 12.2 | 48.0 | 0.0 | 0.0 |
| 18 to 64 years | 51.0 | 53.7 | 68.2 | 66.6 | 63.1 | 29.6 | 53.3 | 33.4 | 66.9 | 39.9 | 52.7 | 54.7 |
| >65 years | 49.0 | 34.7 | 21.7 | 6.9 | 26.5 | 11.0 | 33.5 | 19.1 | 20.9 | 12.1 | 47.3 | 45.3 |
| **Hospitalized with Influenza, n (%)\*** | - | - | - | - | - | - | 290<br>(44.3) | 365<br>(55.7) | 62 494<br>(27.2) | 167 050<br>(72.8) | 4473<br>(47.9) | 4863<br>(52.1) |
| Female sex, % | - | - | - | - | - | - | 54.7 | 52.3 | 63.0 | 53.9 | 6.6 | 4.4 |
| Age, % |  |  |  |  |  |  |  |  |  |  |  |  |
| <18 years | - | - | - | - | - | - | 0.0 | 0.0 | 1.8 | 9.6 | 0.0 | 0.0 |
| 18 to 64 years | - | - | - | - | - | - | 44.9 | 55.9 | 44.4 | 29.3 | 28.0 | 25.3 |
| >65 years | - | - | - | - | - | - | 55.1 | 44.2 | 53.9 | 61.1 | 72.0 | 74.7 |

**Notes:** \* Proportion of obese and non-obese patients among all patients (row percentage); - data not available or below the minimum cell count required (5 individuals)
**Abbreviations:** COVID-19: Coronavirus Disease 2019; CPRD: Clinical Practice Research Datalink; CUIMC: Columbia University Irving Medical Center; SIDIAP:
Information System for Research in Primary Care; STARR-OMOP: Stanford Medicine Research Data Repository; VA-OMOP: United States Department of Veterans Affairs.

The influenza dataset included 4 549 568 patients *diagnosed* and 239 535 patients *hospitalized* with influenza. Among patients *diagnosed* or *hospitalized* with influenza, 688 553 (15%) and 67 257 (28%) were obese, respectively. Aside from VA-OMOP, obesity prevalence was lower among patients *diagnosed* or *hospitalized* with influenza compared to those with COVID-19, with differences ranging from 9% (CPRD) to 18% (IQVIA-OpenClaims) among *diagnosed*, and ranging from 5% (VA-OMOP) to 11% (CUIMC) among *hospitalized* patients.

### Baseline socio-demographics

Overall, *diagnosed* patients were mainly female and aged between 18 and 64 years (Table 1 and Supplementary table 3). However, the proportion of patients aged above 64 years was higher among obese patients with COVID-19 compared to non-obese for SIDIAP, STARR-OMOP, CUIMC, and VA-OMOP and slightly lower for CPRD and IQVIA-OpenClaims. The proportion of patients younger than 18 was very low for obese *diagnosed* COVID-19 patients, with less than 2% in all databases, whereas it was greater than 10% in four databases for obese influenza patients.

Aside from VA-OMOP, in the *hospitalized* cohorts, female sex was more common among obese COVID-19 (ranging from 51% to 55%) and influenza cases (from 55% to 63%) than in non-obese COVID-19 (from 40% to 50%) (Table 1). Overall, *hospitalized* patients were older than those *diagnosed*. In the *hospitalized* cohorts, obese patients with COVID-19 were fairly consistently younger than non-obese (except for SIDIAP) and obese influenza patients. The proportion of patients aged above 65 ranged from 36% to 63% for obese, from 43% to 73% for non-obese and from 54% to 72% for obese influenza patients.

### Baseline comorbidities

In the *diagnosed* cohorts, obese patients with COVID-19 consistently had a higher prevalence of comorbidities compared to non-obese patients (upper part of Figure 1.A and Supplementary table 4). For example, while the prevalence of hypertension for the obese ranged from 30% to 32% in Europe (CPRD and SIDIAP) and from 55% to 81% in the US, in the non-obese it ranged from 12% to 16% and from 26 to 53%, respectively. When compared to obese patients with influenza, obese patients with COVID-19 had a higher prevalence of comorbidities for SIDIAP, STARR-OMOP, and IQVIA-OpenClaims whereas they had fewer comorbidities for CPRD and CUIMC (Figure 1.B, Supplementary table 6).

**Figure 1:**
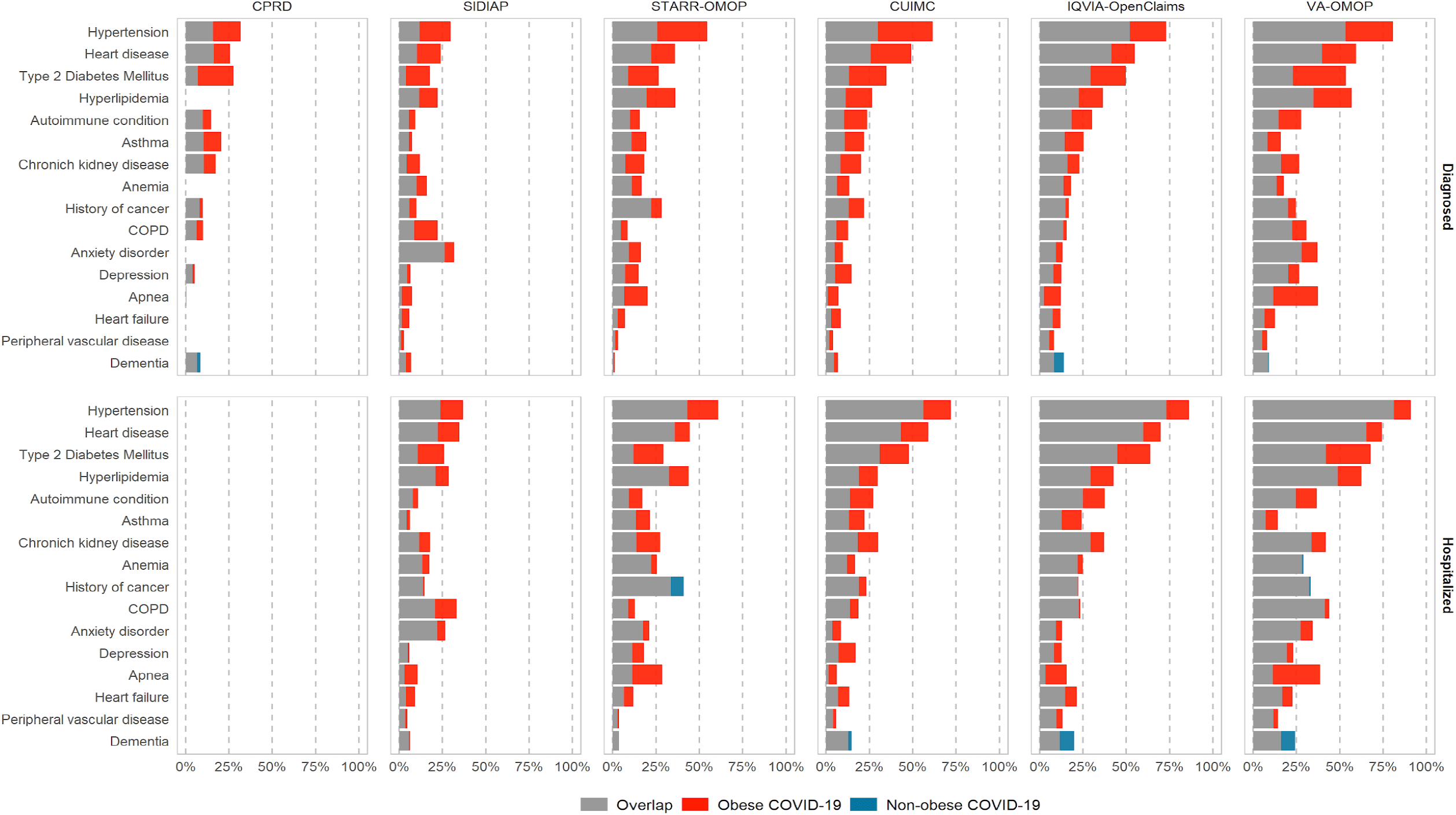
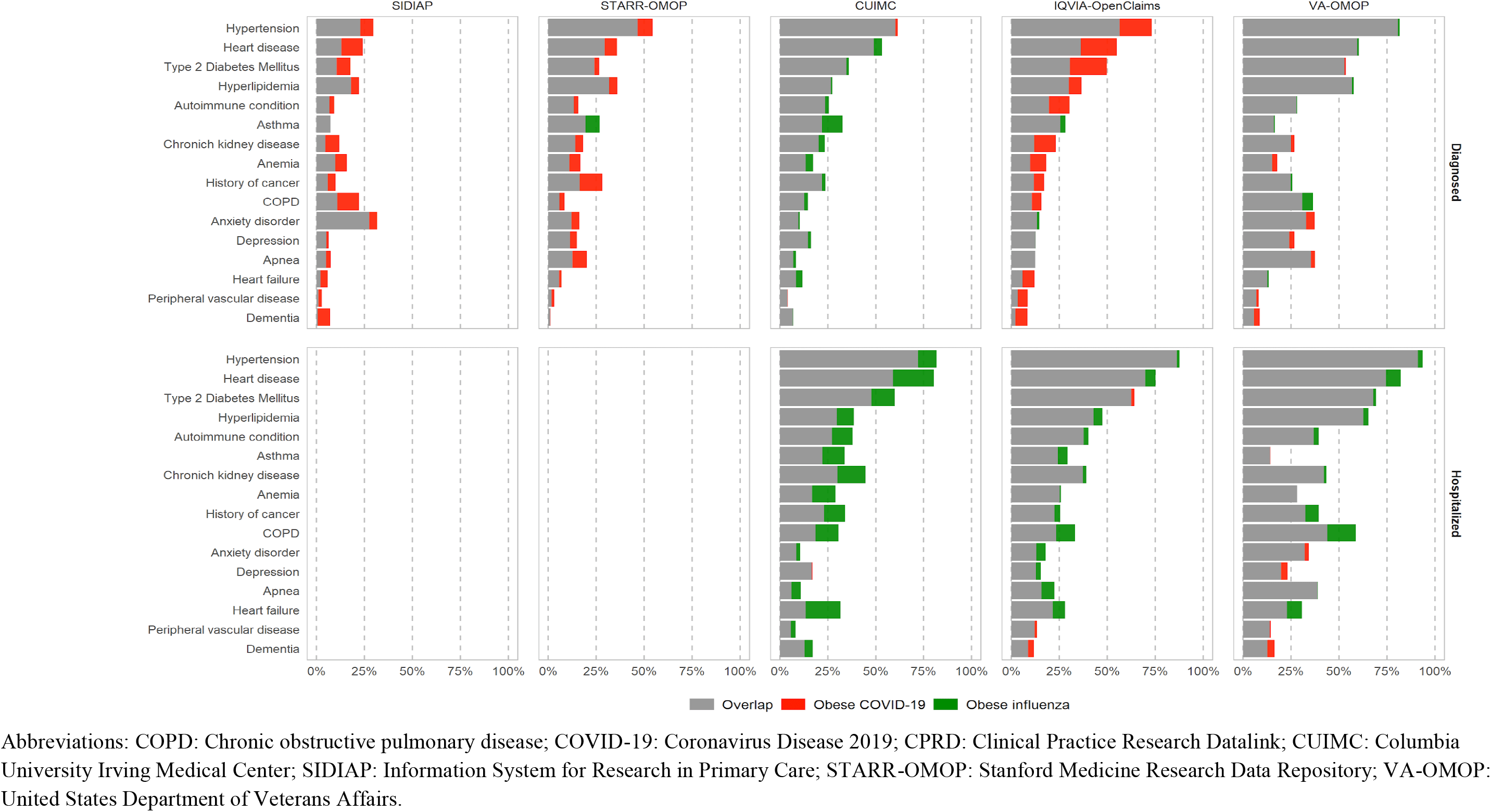
Previous comorbidities: a comparison between obese and non-obese patients with COVID-19 and obese influenza patients. **A. Obese and non-obese COVID-19 patients** Prevalence of comorbidities for obese (red) and non-obese (blue) patients are depicted in overlapped horizontal bars. Grey colour is the overlap between groups (therefore, it shows the lowest value). For example, in CPRD, 32% of obese and 16% of non-obese patients have hypertension. **B. Obese patients with COVID-19 or with Influenza** Prevalence of comorbidities for obese COVID-19 (red) and obese Influenza (green) patients are depicted in overlapped horizontal bars. Grey colour is the overlap between groups (lowest value).

As in the *diagnosed* cohort, *hospitalized* obese patients with COVID-19 had a higher prevalence of comorbidities than non-obese; however, the differences between groups were less obvious (lower part of Figure 1.A., Supplementary table 5). For example, heart disease differed by 20% among those *diagnosed* in VA-OMOP (obese: 60%, non-obese: 40%) and by 9% among those *hospitalized* (obese: 74%, non-obese: 65%). Contrary to the findings for the *diagnosed* cohort, *hospitalized* obese patients with influenza had a higher prevalence of comorbidities compared to COVID-19 patients (lower part of Figure 1.B., Supplementary table 6), except for mental health conditions which were higher in COVID-19 patients in some databases (anxiety in SIDIAP and VA-OMOP and dementia in IQVIA-OpenClaims and VA-OMOP).

### 30-day outcomes of interest

In the *diagnosed* cohorts, hospitalizations rates were higher among obese COVID-19 patients than among non-obese and obese influenza patients in all databases (Figure 2). For example, in IQVIA-OpenClaims, the proportion of patients hospitalized was 32% and 26% for obese and non-obese patients with COVID-19 and 9% for obese patients with influenza. Among obese COVID-19 patients, fatality ranged from 5% to 12%. While CPRD and VA-OMOP had similar fatality in obese and non-obese; in SIDIAP and CUIMC, obese patients had a higher fatality than non-obese (7% vs 3% and 8% vs 5%, respectively). Among obese COVID-19 patients, fatality was higher than among obese influenza patients (range: 0·1% to 3%).

**Figure 2:**
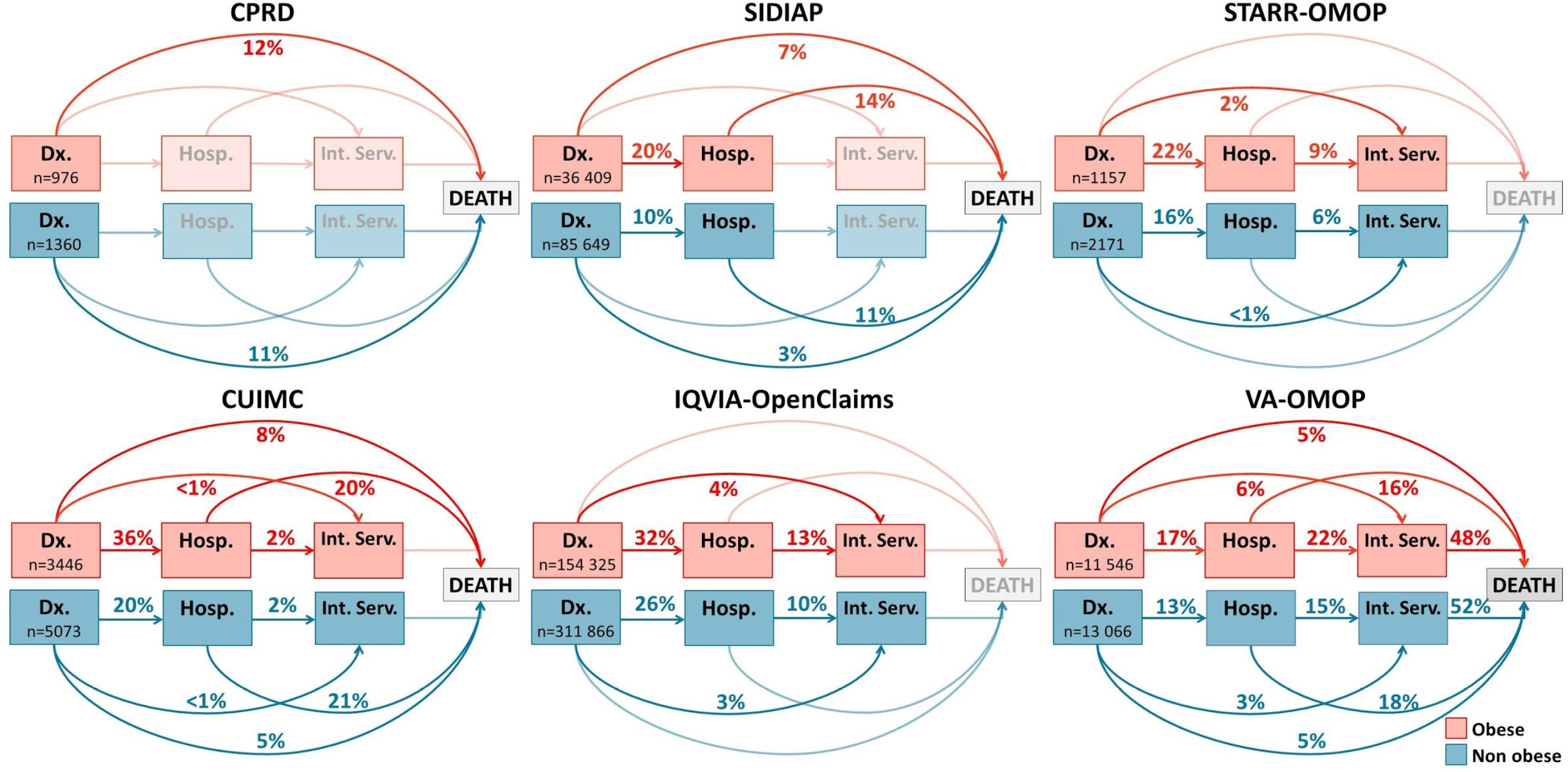
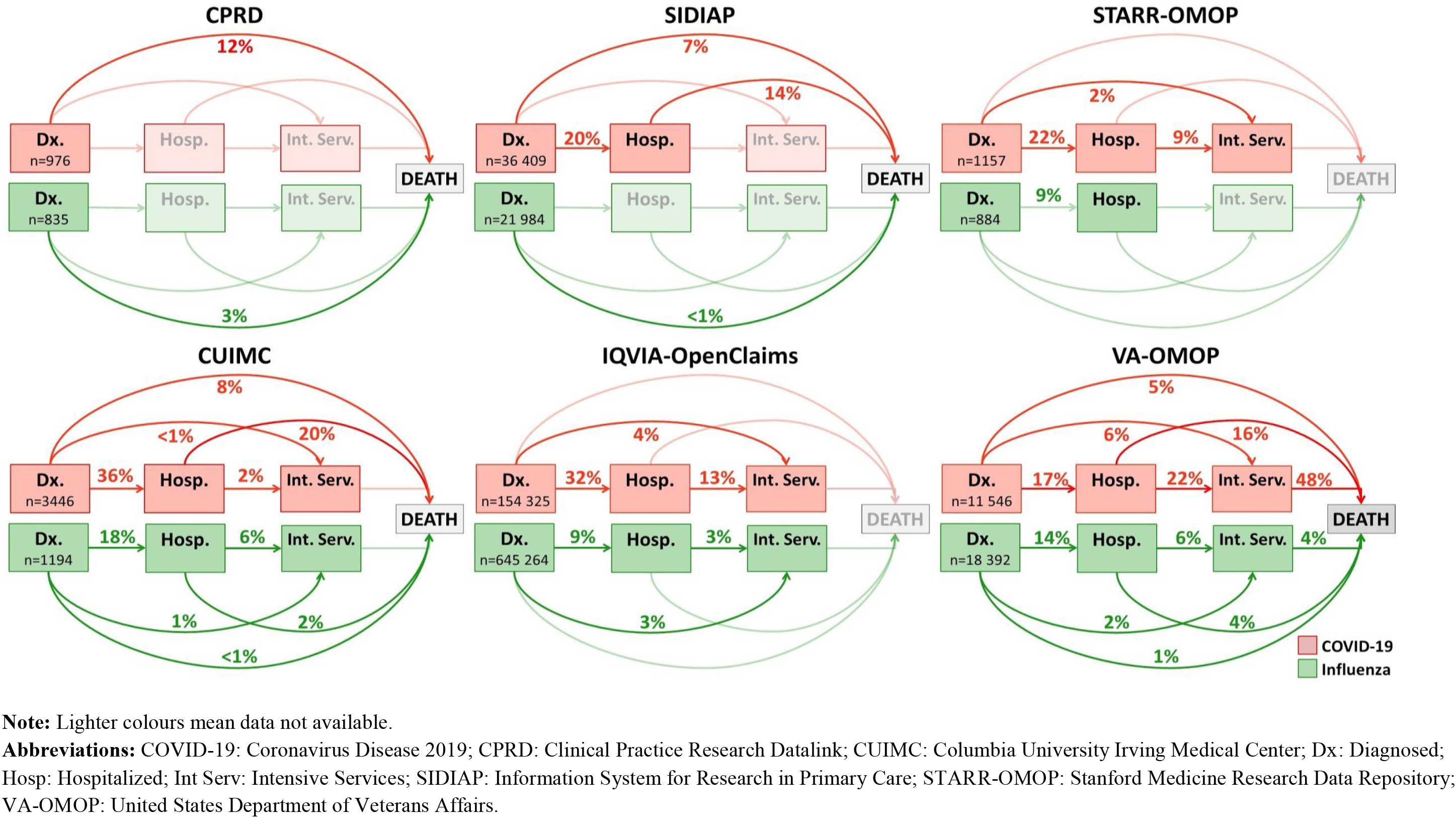
Main outcomes: a comparison between obese and non-obese patients with COVID-19 and obese influenza patients. **A. Obese and non-obese COVID-19 patients** **B. Obese COVID-19 and obese influenza patients**

In the *hospitalized* cohorts, obese patients with COVID-19 required intensive services more frequently than non-obese in STARR-OMOP (obese: 9% vs non-obese: 6%), IQVIA-OpenClaims (13% vs 10%) and VA-OMOP (22% vs 15%). Percentages in CUIMC were too small to assess differences (2·3% vs 2·0%). *Hospitalized* obese patients with COVID-19 also required intensive services more often than obese patients with influenza in IQVIA-OpenClaims (13% vs 3%) and VA-OMOP (22% vs 6%) whereas in CUIMC (2% vs 6%) the opposite was observed.

There were no notable differences in fatality between obese and non-obese *hospitalized* patients with COVID-19: 14% vs 11% in SIDIAP, 20% vs 21% in CUIMC and 16% vs 18% in VA-OMOP. However, fatality among obese COVID-19 cases was much higher than among obese influenza patients (2% in CUIMC and 4% in VA-OMOP).

Overall, obese patients with COVID-19 had adverse events occurring in the 30 days after the index date more frequently than non-obese (Table 2). This was especially evident for the most frequent events, namely acute respiratory distress syndrome (ARDS) (range: 15% to 46% among obese, range: 10% to 41% among non-obese) and heart failure (7% to 23% among obese, 3% to 17% among non-obese) during hospitalization. Sepsis, cardiac arrhythmia, and total cardiovascular disease events were also slightly more frequent among obese COVID-19 patients compared to non-obese. Acute kidney injury was the only outcome that was more frequent among non-obese; however, except for CUIMC, differences were small. For the rest of the reported events, numbers were too small to assess differences between obese and non-obese COVID-19 patients.

**Table 2:** 30-day events in obese and non-obese patients hospitalized with COVID-19.

|  | STARR-OMOP |  | CUIMC |  | IQVIA-OpenClaims |  | VA-OMOP |  |
| --- | --- | --- | --- | --- | --- | --- | --- | --- |
|  | Obese | Non-obese | Obese | Non-obese | Obese | Non-obese | Obese | Non-obese |
| **n** | 274 | 341 | 1408 | 1192 | 50 863 | 8228 | 2918 | 2592 |
| **Cardiovascular events, %** |  |  |  |  |  |  |  |  |
| Acute myocardial infarction | - | - | 1.3 | 1.1 | 2.1 | 2.2 | 5.5 | 6.9 |
| Cardiac arrhythmia during hospitalization | 16.8 | 14.1 | 9.8 | 9.3 | 13.1 | 11.7 | 33.4 | 31.4 |
| Heart failure during hospitalization | 15.7 | 9.1 | 6.5 | 3.1 | 7.0 | 5.1 | 22.9 | 17.4 |
| Stroke (ischemic or hemorrhagic) | - | - | 2.6 | 2.5 | 1.7 | 1.9 | 2.4 | 4.0 |
| Total cardiovascular disease events | 11.7 | 8.8 | 7.3 | 5.9 | 8.2 | 7.6 | 20.6 | 19.0 |
| **Thromboembolic events, %** |  |  |  |  |  |  |  |  |
| Deep vein thrombosis | - | - | 2.1 | 1.8 | 2.2 | 1.7 | 4.9 | 3.8 |
| Pulmonary embolism | - | - | 2.2 | 1.8 | 1.8 | 1.5 | 3.8 | 3.3 |
| **Other events %** |  |  |  |  |  |  |  |  |
| Acute kidney injury | 8.4 | 9.1 | 18.1 | 25.4 | 11.1 | 10.4 | 17.1 | 18.6 |
| Acute pancreatitis | - | - | - | - | 0.2 | 0.2 | 0.4 | 0.7 |
| ARDS during hospitalization | 14.6 | 10.0 | 17.9 | 15.2 | 34.9 | 30.5 | 46.2 | 41 |
| Dialysis | - | - | - | - | 4.3 | 3.2 | 8.3 | 5.9 |
| Gastrointestinal bleeding | - | - | 1.7 | 1.0 | 1.3 | 1.3 | 3.1 | 4.7 |
| Hepatic failure | - | - | 0.8 | - | 0.2 | 0.2 | 1.6 | 0.8 |
| Sepsis during hospitalization | 9.5 | 7.9 | 5.6 | 6.0 | 17.4 | 16.2 | 24.3 | 22.0 |

**Notes:** - data not available or below the minimum cell count required (5 individuals)
**Abbreviations:** ARDS: Acute Respiratory Distress Syndrome; CUIMC: Columbia University Irving Medical Center; STARR-OMOP: Stanford Medicine Research Data Repository; VA-OMOP: United States Department of Veterans Affairs.

## Discussion

In this large cohort including 627 044 COVID-19 and 4 549 568 influenza patients from the US, Spain, and the UK, we found that the prevalence of obesity was higher among COVID-19 patients *hospitalized* compared to those *diagnosed* and was higher in both COVID-19 cohorts compared to the influenza cohorts. Obese patients *diagnosed* and *hospitalized* with COVID-19 were more commonly female, and presented with more comorbidities and adverse outcomes than non-obese patients. Although obese COVID-19 patients were younger and less likely to have comorbidities than obese influenza patients in the *hospitalized* cohorts, they more frequently had adverse outcomes.

Given the prevalence of obesity in the US (37%), the UK (27%) and Spain (24%), a high proportion of obese patients among COVID-19 cases was expected.^22^ However, the prevalence of obesity among COVID-19 patients was higher than the general population in the three countries, which is suggestive of an increased risk of diagnosis in obese patients. Obese individuals could be more likely to seek care and be tested for SARS-CoV-2 since they are a high-risk, multimorbid population and could be more prone to respiratory symptoms due to their compromised pulmonary function. Alternatively, they might have an increased risk of COVID-19 infection that could be explained through differences in exposure and/or vulnerability. Two studies from Spain and the UK found a higher risk of being diagnosed with COVID-19 in obese individuals.^7,23^ We also found a higher proportion of obesity among *hospitalized* COVID-19 patients compared to those *diagnosed*. The prevalence of obesity from this study was similar to three cohort studies from the US that reported 40%, 42% and 48% of hospitalized patients were obese.^9-11^ These findings could reflect a preferential care in high-risk populations but also an increased risk of severe disease among obese patients.

Prior characterization studies from the hospital setting have consistently reported that COVID-19 hospitalized patients are more frequently male.^10,11^ We found, however, that women predominate among hospitalized obese patients. Since the prevalence of obesity is similar for men and women in the three countries, our findings could reflect that obesity is a greater risk factor for hospitalization among women compared to men.^22^ Some of the biological hypotheses that have been posited to explain the greater risk of poor COVID-19 outcomes among men are similar to those proposed for obese individuals. For example, one hypothesis argues that sex differences in the immune response increase men’s vulnerability. Men have higher levels of interleukin-6 and tumour necrosis factor alpha, which are the main drivers of the phenomenon of “cytokine storm” that has been observed in severe COVID-19 patients.^24,25^ Obese individuals also have higher levels of these pro-inflammatory cytokines.^26^ Similarly, men have higher levels of angiotensin-converting enzyme 2, which are also elevated in obese individuals.^25,27^

We also found that compared to non-obese, patients with obesity were older among those *diagnosed* with COVID-19 and younger in those *hospitalized*. The former observation might be explained by the early identification of both older age and obesity as risk factors for COVID-19.^4^ This could have increased the frequency of testing among people with these characteristics. Conversely, the differences observed in the hospital setting could be related to a greater risk of severe outcomes among individuals with obesity younger than 60.^28^ Although younger individuals have less risk of infections and complications than older people due to having fewer comorbidities and a stronger immune system,^29,30^ this is not the case for obese individuals.^31,32^ Therefore, the more fragile health of obese patients even in the young may contribute to the observed age pattern for the individuals hospitalized with obesity.

Obese individuals differed also in terms of comorbidities. Unsurprisingly, the highest differences were observed in obesity-related conditions, such as hypertension, diabetes, and heart disease, which have been identified as risk factors for severe COVID-19 outcomes.^7,8,10,13^ The selected comorbidities likely represent only a small proportion of the differences that should be considered when assessing the links between obesity and COVID-19. For instance, obesity is strongly interlinked with socioeconomic status and ethnicity, and disproportionately affects disadvantaged populations.^29^ In the US, the prevalence of obesity is higher among Hispanic and African Americans than among their White counterparts. These disparities have been hypothesized as a contributing factor explaining ethnic differences in the proportion of hospitalizations.^33^ Given the links between obesity and the social determinants of health, obese individuals might be more at risk of COVID-19 infection and complications due to more precarious working conditions and barriers in healthcare access. Unfortunately, due to data unavailability, we could not directly explore socioeconomic status or ethnicity differences in our study.

Finally, obese individuals experienced more frequently adverse events than non-obese, including hospitalization and requirement of intensive services. These results, however, must be interpreted carefully considering the differences in socio-demographics and comorbidities between obese and non-obese individuals. Despite this limitation, our results are in line with previous studies where obesity was associated with an increased risk of hospitalization and mortality, adjusting for sex, age, and comorbidities.^7,8,10,13^ Given the scarcity of evidence regarding the frequency of specific adverse events during hospitalization among obese patients, our findings are of special interest to the field and should be addressed in upcoming associative studies.

Since obesity has been associated with different forms of influenza, an association with COVID-19 was expected from the beginning of the pandemic.^34^ Interestingly, we found that the prevalence of obesity was higher among both COVID-19 *diagnosed* and *hospitalized* patients compared to those with seasonal influenza, which may be suggestive of an inherent vulnerability towards COVID-19 among obese patients. In both COVID-19 and influenza cohorts, female sex was more frequent than male sex. A large study comparing hospitalized patients with COVID-19 to those with influenza found that COVID-19 patients were predominantly male whereas those with influenza were mostly women.^35^ While our study replicated those findings for non-obese patients, we observed that women predominated among obese patients hospitalized with COVID-19. Thus, our results provide a finer picture of the most frequent sex of patients with COVID-19 among those with obesity. Older age has been associated with an increased risk of more severe forms of both COVID-19 and influenza,^3^ however, we observed obese patients with COVID-19 were younger than those with influenza. Our findings are in line with previous studies that reported younger ages among individuals with H1N1 and COVID-19 compared to seasonal influenza (obese and non-obese individuals together).^35,36^ Finally, although obese patients diagnosed with COVID-19 had more comorbidities than obese influenza patients the opposite was observed among hospitalized patients. This could reflect either a higher virulence of SARS-CoV-2 compared to influenza or a differential pattern in clinical practice, with a lower threshold for hospitalization in COVID-19 cases. However, obese patients with COVID-19 consistently had worse outcomes than influenza patients. These results put in to question the justification that worse outcomes among obese patients with COVID-19 are merely due to a higher prevalence of co-existing conditions.^37^

Our study has several strengths, such as its large amount of data. This study contains information on 627 044 COVID-19 cases from six databases of three different countries and provides a wide overview of the characteristics and outcomes of patients with and without obesity. Almost 29 000 unique aggregate characteristics were generated and made publicly available. This was accomplished through the coordinated efforts of the OHDSI community to provide a rapid response to the COVID-19 pandemic. We employed a federated analysis approach that allowed us to respect the confidentiality of patient records at all times, and we emphasized transparency throughout the study, making methods, tools, and results publicly available.

Our study also has limitations. One limitation is its descriptive nature. Since we aimed to characterize and compare patients with and without obesity, statistical tests and modelling were out of scope in the developed analytical packages. However, our findings were consistent across databases. In addition, the exhaustive characterization performed in this study supports the generation of new hypotheses that can be tested in detail in future studies. Second, we cannot exclude a selection bias of COVID-19 cases due to underreporting in the context of testing restrictions and asymptomatic or paucisymptomatic cases that usually do not seek medical care. Additionally, testing policies have varied across countries and time depending on the course of the pandemic. The inclusion of COVID-19 cases clinically diagnosed (without being tested) in different countries and settings likely provided consistency to our data, although it might have incurred in false positives. Third, we did not have information on BMI as a continuous variable, which prevented us from investigating the impact of different categories of obesity in COVID-19 outcomes. This might explain the higher proportion of comorbidities and outcomes observed in the US databases, as obese individuals from the US might be more obese than those from Europe. Fourthly, we cannot discard that the differences found in the COVID-19/seasonal influenza comparison may have been influenced by temporal changes in clinical practice standards and coding. Further, the use of influenza vaccination among high-risk population groups likely contributed to the observed low proportion of adverse events among influenza patients.^38^ Finally, this study was underpinned by routinely-collected data which can raise concerns about the quality of the data. Some databases are prone to oversampling certain groups of people as a result of how these data are captured (e.g. the Veterans Affairs system historically serves more men than women, routine claims data may only reflect health outcomes in commercially insured populations, etc). Obesity, comorbidities, and outcomes were assessed based on having a record of a condition/measurement, therefore they may be underestimated. Even still, the consistency of our findings across several databases that differ by setting and country lends credence to the generalizability of our findings.

In this large international cohort, we showed that obese patients with COVID-19 were more likely to be female, had more comorbidities and worse outcomes than non-obese patients. We provide novel evidence that the prevalence of obesity is higher among COVID-19 patients compared to those with seasonal influenza and that obese hospitalized COVID-19 patients have worse outcomes than obese patients with influenza, despite presenting with fewer comorbidities. Our results may be useful in guiding clinical practice and aid future preventative strategies for obese individuals, as well as providing useful data to support subsequent association studies focussed on obesity and COVID-19.

## Data Availability

Analyses were performed locally in compliance with all applicable data privacy laws. Although the underlying data is not readily available to be shared, authors contributing to this paper have direct access to the data sources used in this study. All results (e.g. aggregate statistics, not presented at a patient-level with redactions for minimum cell count) are available for public inquiry. These results are inclusive of site-identifiers by contributing data sources to enable interrogation of each contributing site. All analytic code and result sets are made available at: https://github.com/ohdsi-studies/Covid19CharacterizationCharybdis

https://github.com/ohdsi-studies/Covid19CharacterizationCharybdis

## Ethical approval

All the data partners received Institutional Review Board (IRB) approval or exemption. STARR-OMOP had approval from IRB Panel #8 (RB-53248) registered to Leland Stanford Junior University under the Stanford Human Research Protection Program (HRPP). The use of VA data was reviewed by the Department of Veterans Affairs Central IRB, was determined to meet the criteria for exemption under Exemption Category 4(3), and approved for Waiver of HIPAA Authorization. The research was approved by the Columbia University Institutional Review Board as an OHDSI network study. The use of SIDIAP was approved by the Clinical Research Ethics Committee of the IDIAPJGol (project code: 20/070-PCV). The use of CPRD was approved by the Independent Scientific Advisory Committee (ISAC) (protocol number 20_059RA2). The use of IQVIA-OpenClaims was exempted from IRB approval.

## Transparency declaration

Lead authors affirm that the manuscript is an honest, accurate, and transparent account of the study being reported; that no important aspects of the study have been omitted; and that any discrepancies from the study as planned have been explained.

## Contributorship statement

MR, ER, APU, PR, DPA, KK and TDS conceived and designed the study. SLD, TF, KEL, MEM, KN, JDP, CGR, NHS, PR, KK and TDS coordinated data contributions at their respective sites. AP, AGS, TF, SFB, JDP, KK and TDS analyzed the data; MR, ER and AP produced the figures and tables. MR, ER, EB, DRM, FN, PR, LMS, DPA, KK and TDS interpreted the data. MR, ER and TDS searched the literature and wrote the first draft with insightful contributions from EB, LYHL, JCEL, DRM, FN, PR, LMS, DPA and KK. All authors contributed to the revision of the first draft, reviewed and approved the final version of the manuscript.

## Acknowledgements

We would like to acknowledge the patients who suffered from or died of this devastating disease, and their families and carers. We would also like to thank the healthcare professionals involved in the management of COVID-19 during these challenging times, from primary care to intensive care units.

## Data sharing statement

**Abbreviations**
ARDS: Acute Respiratory Distress Syndrome
BMI: Body Mass Index
CDM: Common Data Model
COVID-19: Coronavirus disease 2019
CPRD: Clinical Practice Research Datalink
CUIMC: Columbia University Irving Medical Center
EHR: Electronic Health Record
IRB: Institutional Review Board
OHDSI: Observational Health Data Sciences and Informatics
OMOP: Observational Medical Outcomes Partnership
SARS-CoV-2: severe acute respiratory syndrome coronavirus 2
SIDIAP: Information System for Research in Primary Care
SNOMED: Systematized Nomenclature of Medicine
STARR-OMOP: Stanford Medicine Research Data Repository
UK: United Kingdom
US: United States
VA-OMOP: United States Department of Veterans Affairs
WHO: World Health Organization

